# Comparison of length of liver among patients with and without fatty change in liver

**DOI:** 10.1101/2025.01.11.25320400

**Authors:** Shahriar Muttakin, Azmal Kabir Sarker, Farzana Rahman Adrita, Nafisa Kaisar Tamanna

## Abstract

**Objective:** The retrospective cross-sectional study was done to assess the change of length of liver across the grades of fatty change, across the ages of individual as well as between the male and female sexes.

**Methods:** The liver length measurement and qualitative visual grading of fatty change (from grade 0 to grade 3), obtained from patients who underwent ultrasound of liver at our institute between July and October of 2024 was retrospectively retrieved. The age of patient, length of liver and the grading of fatty change was used as the variables for the analyses.

**Results:** The mean lengths of liver were found to be significantly increasing with increased grades fatty change (grade 0, grade I and grade II), both in males and females (p < 0.01). Male patients were affected by fatty change in liver (both grades I and II), more in comparison to the females (Pearson’s Chi-squared value 4.53, p = 0.033). Being at older age is significantly associated with being affected by fatty change in liver (both grades I and II) in females (p<0.01). Liver with fatty changes (grade I and II combined) were significantly larger than those without fatty change both in males as well as in females (p < 0.01). A weak positive association of length of liver and advancing age was found only among females without fatty change (R 0.29, p = 0.046).

**Conclusions:** Findings from this study is expected to have implications in the clinical and lifestyle management of NAFLD and future research

## INTRODUCTION

Transabdominal ultrasound is a recommended imaging method for the screening of non-alcoholic fatty liver disease (NAFLD). (1). Visual comparison of echogenicity between liver parenchyma and the cortex of the right kidney is used for the qualitative assessment for fatty change in liver or hepatic steatosis (2). The frequent finding of fatty change in liver in the practice of medical diagnostic transabdominal ultrasound is attributable to the prevalence of NAFLD being 33.9% in Bangladeshi population (3). This is higher than the global prevalence of 25.2% (4) while the reported risk factors of NAFLD in Bangladesh are increasing age, obesity, diabetes, and hypertension (3).

The liver size is reportedly larger in patients with NAFLD with progressive enlargement with increasing grades of fatty change (5). To verify this relationship among liver size, patients’ age and grade of fatty change, this study was conducted with an aim to assess the change of length of liver across the grades of fatty change, across the ages of individual as well as between the male and female sexes.

## PATIENTS AND METHODS

The study was retrospectively conducted using the liver measurement data and qualitative visual grading of fatty change, obtained from patients who underwent ultrasound of liver at our institute between July and October of 2024. Patients with focal parenchymal lesion, coarse hepatic parenchyma or acute parenchymal disease were excluded.

In all patients, the length of liver was measured in two-dimensional images obtained in the right anterior axillary line while placing the electronic calipers between dome of the diaphragm and the tip of the liver (6, 7). The qualitative classification of fatty change in liver or hepatic steatosis was done following the grading reported by Saadeh et al and mentioned in box 1 (2).

### Box 1

Grading of fatty change in liver

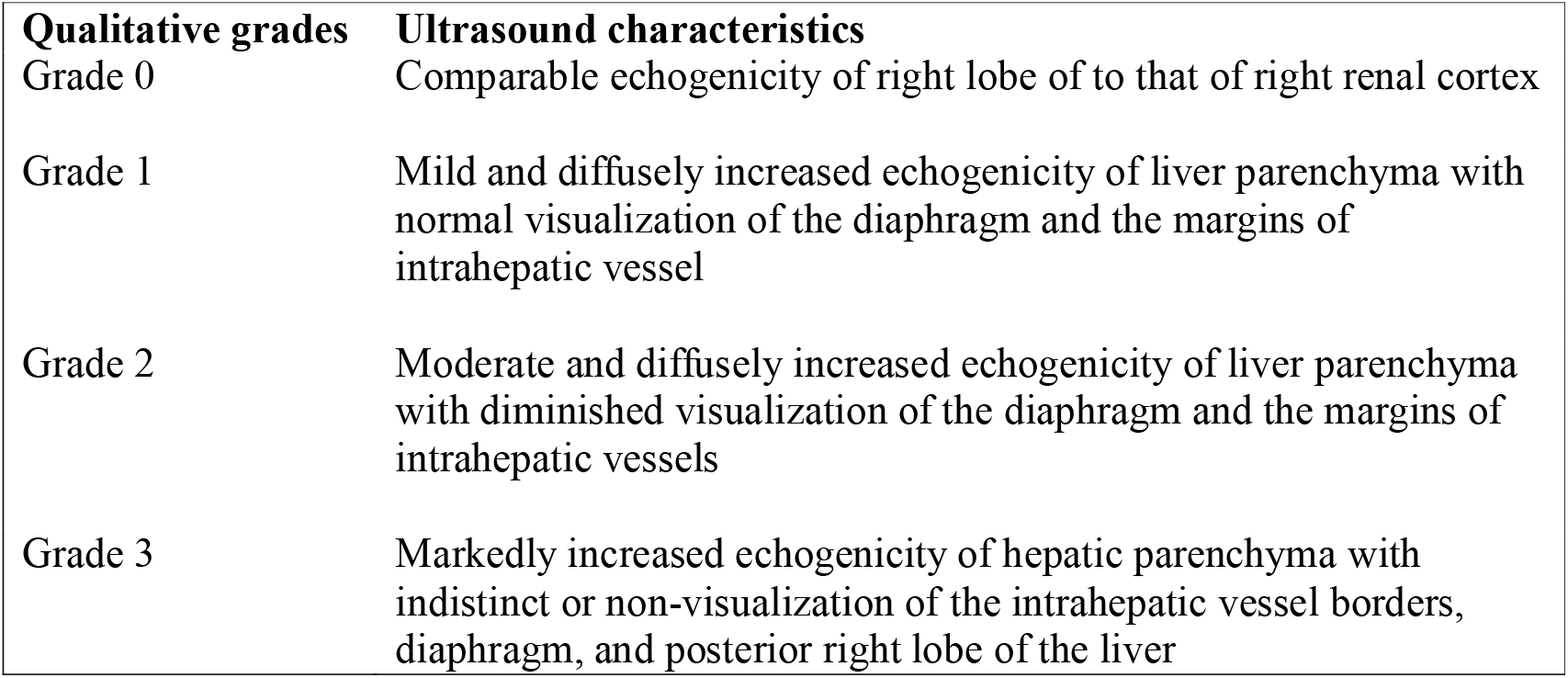

**Box 1: Grading of fatty change in liver**
| **Qualitative grades** | **Ultrasound characteristics** |
| --- | --- |
| Grade 0 | Comparable echogenicity of right lobe of to that of right renal cortex |
| Grade 1 | Mild and diffusely increased echogenicity of liver parenchyma with normal visualization of the diaphragm and the margins of intrahepatic vessel |
| Grade 2 | Moderate and diffusely increased echogenicity of liver parenchyma with diminished visualization of the diaphragm and the margins of intrahepatic vessels |
| Grade 3 | Markedly increased echogenicity of hepatic parenchyma with indistinct or non-visualization of the intrahepatic vessel borders, diaphragm, and posterior right lobe of the liver |

All statistical tests were done on R. The box-violin plots were generated using the “ggstatsplot”, and Correlation plots were generated using the “ggplot2” functions.

## RESULTS

The analysis was conducted on 217 patients. There were 114 (52.5%) males with mean (±SD) age of 42.1 ±14.2 years and 103 (47.5 %) females with mean (±SD) age of 37.3 ±13.9 years. No patient was classified to have grade 3 fatty change. The mean age of patients (both sexes combined) having grade I fatty change was found to be significantly different from those without fatty change (p<0.01). However, the difference of age was no longer significant when further analysis was done within males and females (table 1). The mean lengths of liver were found to be significantly different among the three categories of fatty change (grade 0, grade I and grade II), in males, females as well as when analyzed both sexes combined (table 1 and figure 1).

**Table 1:**
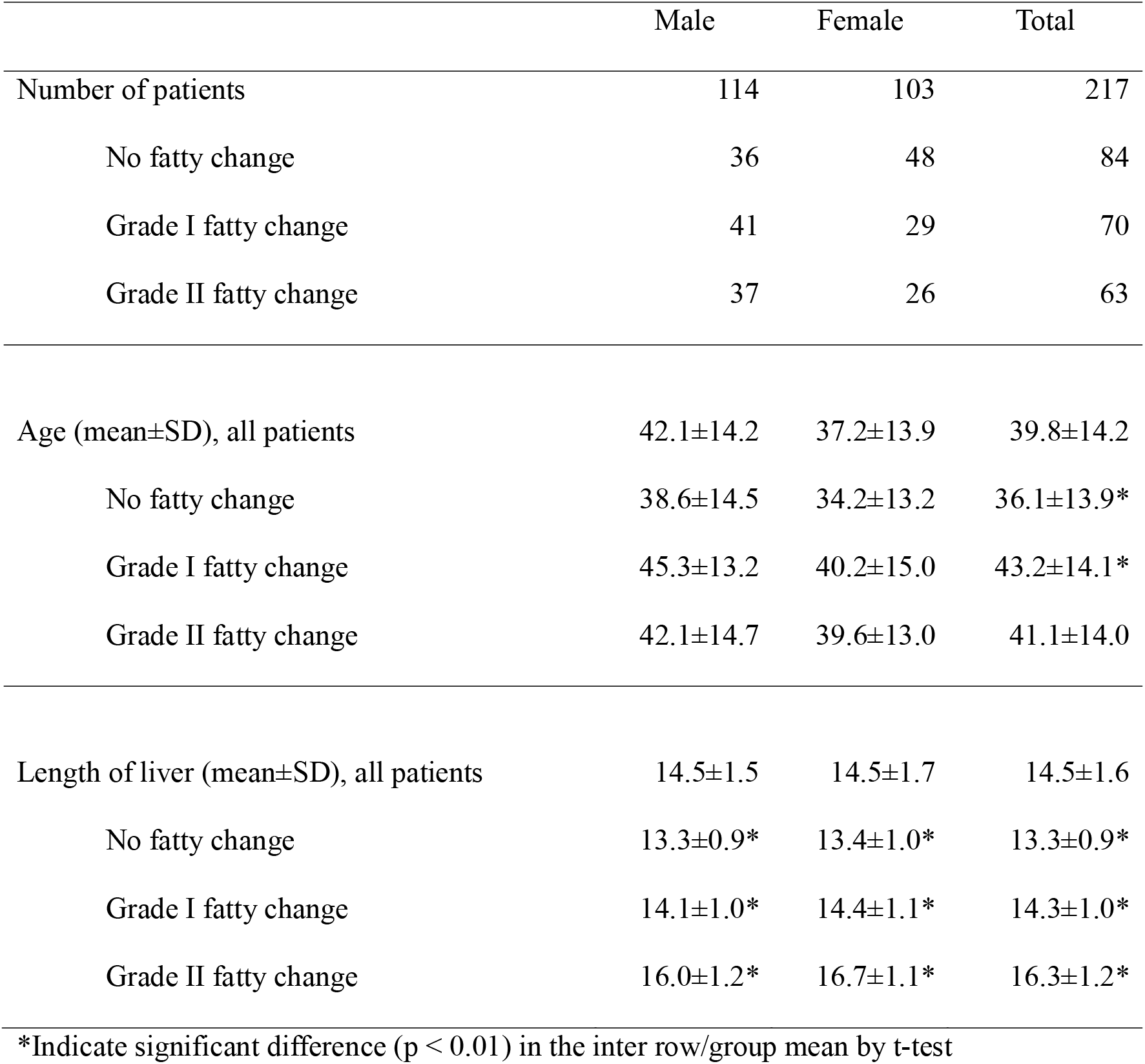
Characteristic of study patients.

|  | Male | Female | Total |
| --- | --- | --- | --- |
| Number of patients | 114 | 103 | 217 |
| No fatty change | 36 | 48 | 84 |
| Grade I fatty change | 41 | 29 | 70 |
| Grade II fatty change | 37 | 26 | 63 |
| Age (mean±SD), all patients | 42.1±14.2 | 37.2±13.9 | 39.8±14.2 |
| No fatty change | 38.6±14.5 | 34.2±13.2 | 36.1±13.9* |
| Grade I fatty change | 45.3±13.2 | 40.2±15.0 | 43.2±14.1* |
| Grade II fatty change | 42.1±14.7 | 39.6±13.0 | 41.1±14.0 |
| Length of liver (mean±SD), all patients | 14.5±1.5 | 14.5±1.7 | 14.5±1.6 |
| No fatty change | 13.3±0.9* | 13.4±1.0* | 13.3±0.9* |
| Grade I fatty change | 14.1±1.0* | 14.4±1.1* | 14.3±1.0* |
| Grade II fatty change | 16.0±1.2* | 16.7±1.1* | 16.3±1.2* |

\*Indicate significant difference ( $p < 0.01$ ) in the inter row/group mean by t-test

**Figure 1:**
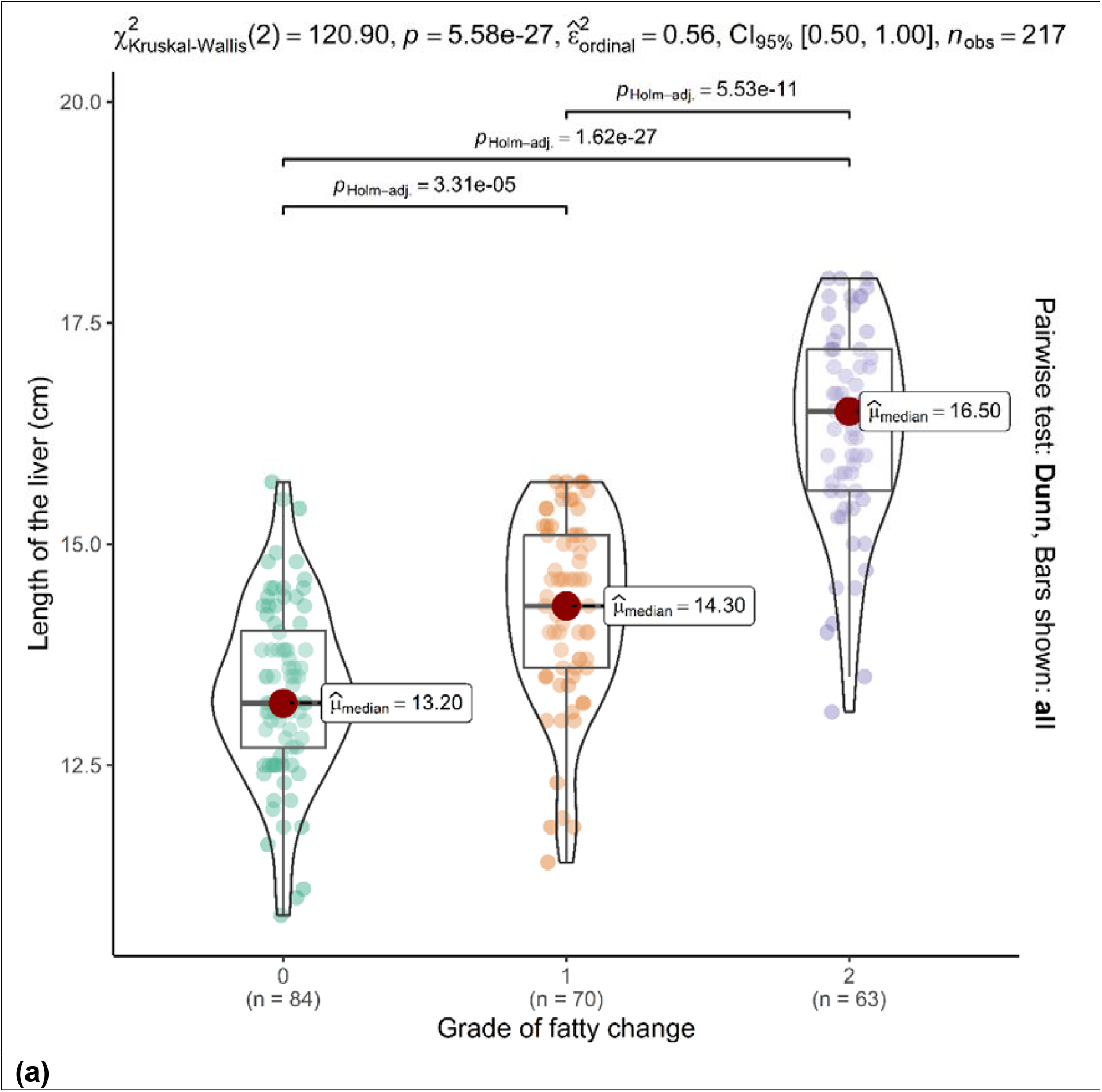
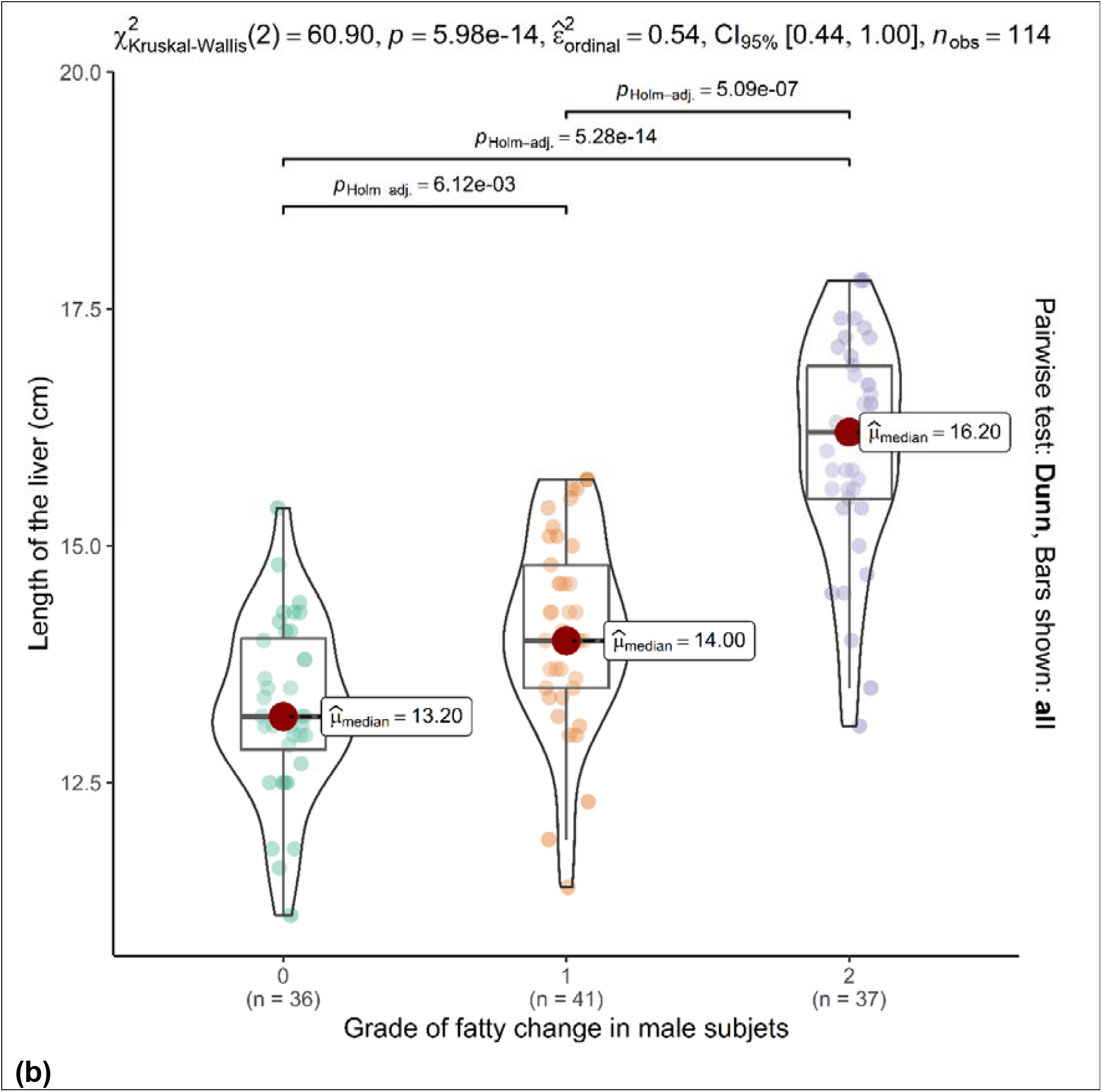
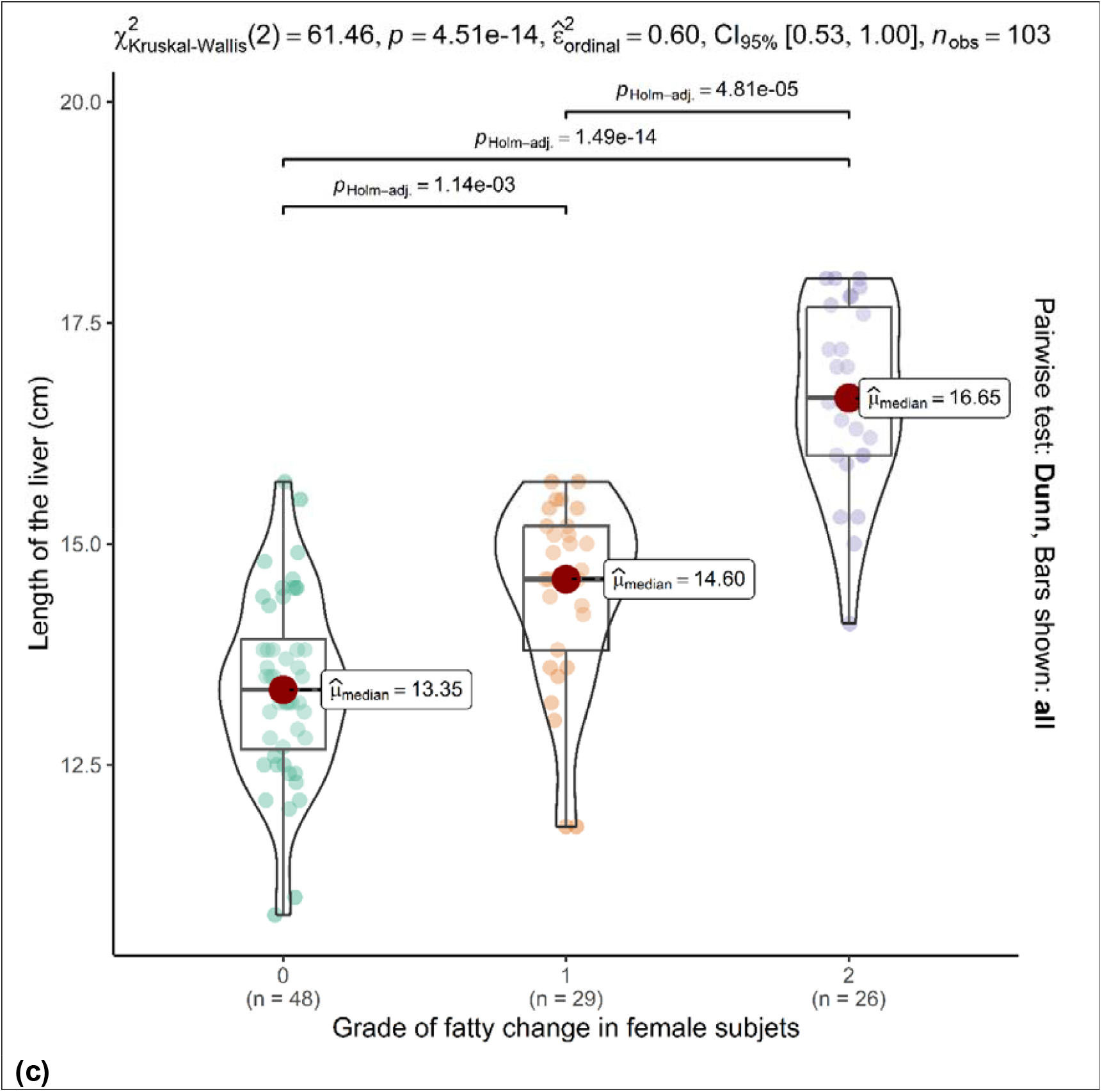
(a) Length of liver in Y-axis plotted against grade of fatty liver in X-axis, in all patients, n = 217, (b) in the male patients, n = 114, (c) in the female patients, n = 103. The grade 0 means no fatty change. The mean values are significantly different with the Holm-adjusted p-value being < 0.01.

We found grade 0 fatty change in liver among 48 of the 103 female patients and among 36 of 114 male patients. Subsequently, a Chi square test revealed significant association of male sex with the state of being affected by fatty change in liver (both grade I and II combined) in our study patients. The Pearson’s Chi-squared value was 4.53 with p-value being 0.033.

A significant association was found of being at older age (both sexes combined) with being affected by fatty change in liver (both grades combined) in our study patients. The mean (± SD) age of patients without fatty change liver was 36.1 ± 13.9 years while the age of patients with fatty changes (grade I and II combined) was 42.2 ± 14 years (p < 0.01). However, when further analyzed within male and female patients, the difference of age was significant only among the females but not in the males (table 2).

**Table 2:** Difference of age and liver length in patients without and with (both grades combined) fatty change.

|  | **Without fatty change**<br>**(grade 0)** | **Fatty change**<br>**(grade I and II)** | **t-test p-value** |
| --- | --- | --- | --- |
| **All patients** | (n=84) | (n=133) |  |
| Age | 36.1 ± 13.9 | 42.2 ± 14 | < 0.01 |
| Length of liver | 13.3±0.9 | 15.2±1.5 | <0.01 |
| **Male** | (n=36) | (n=78) |  |
| Age | 38.6±14.5 | 43.8±13.9 | 0.08 |
| Length of liver | 13.3±0.9 | 15.0±1.5 | <0.01 |
| **Female** | (n=48) | (n=55) |  |
| Age | 34.2±13.2 | 39.9±14.0 | 0.04 |
| Length of liver | 13.4±1.0 | 15.5±1.5 | <0.01 |

Liver with fatty changes (grade I and II combined) were significantly larger than those without fatty change in all patients (both sexes combined). The mean (±SD) length of liver without fatty change was 13.3 ± 0.98 which was clearly in contrast to those with fatty change 15.2 ± 1.49 (p < 0.01). Similar difference was seen when further analyzed within male and female patients (table 2). While checking for the correlation between length of liver with age, a weak positive association was found only among the females without fatty change (table 3, figure 2).

**Table 3:** Correlation of age with length of liver.

| Dataset | Pearson correlation coefficient (R) | p-value |
| --- | --- | --- |
| All patients (n= 217) | 0.06 | 0.38 |
| All patients with no fatty change (n=84) | 0.08 | 0.49 |
| All patients with grade I fatty change (n=70) | -0.13 | 0.29 |
| All patients with grade II fatty change (n=63) | -0.11 | 0.39 |
| All male patients (n=114) | 0.03 | 0.78 |
| Male with no fatty change (n=36) | -0.19 | 0.26 |
| Male with grade I fatty change (n=41) | 0.07 | 0.65 |
| Male with grade II fatty change (n=37) | -0.19 | 0.26 |
| All female patients (n=103) | 0.15 | 0.12 |
| Female with no fatty change (n=48) | 0.29 | 0.046* |
| Female with grade I fatty change (n=29) | -0.15 | 0.44 |
| Female with grade II fatty change (n=26) | -0.14 | 0.48 |

\*Indicates significant difference ( $p < 0.05$ )

**Figure 2:**
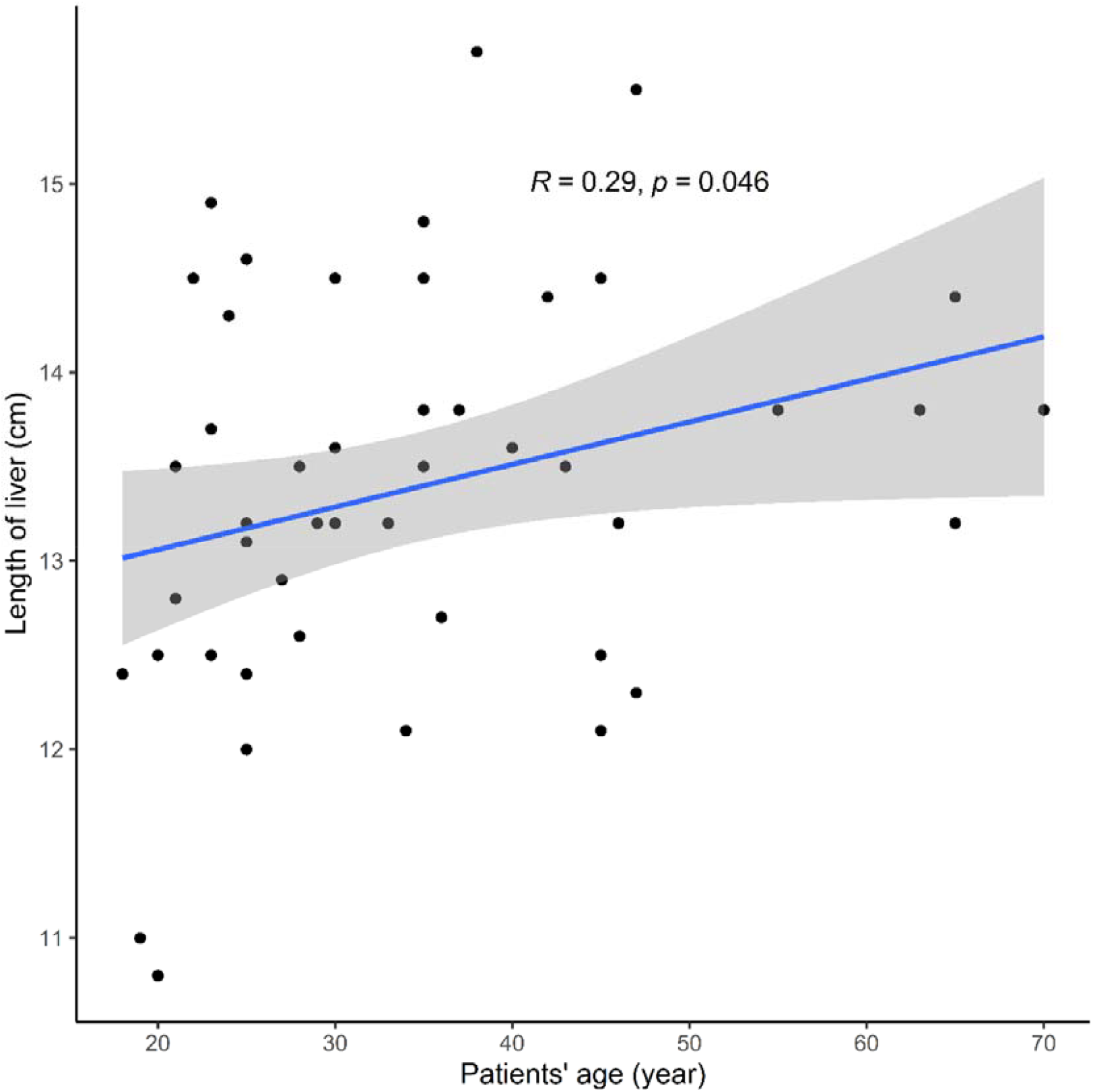
Age versus length of liver in females (n = 48) with no fatty change in liver; the subset among all others shows significant correlation of length with increasing age.

## DISCUSSION

We observed increasing liver length across increasing grades of fatty change. This finding matches with a recent report (5). In our study the age of male patients without fatty change weren’t different from those with fatty change. This is contradictory to the known lower prevalence of NAFLD in males younger than 40 in comparison to males ages 40 and older (8). The female patient without fatty change in this study were significantly younger than those who had fatty change. This is explainable partly by the previous observation of women being affected by NAFLD around their menopause (8).

## CONCLUSION

Our study confirms enlargement of liver across the increasing grade of fatty change while adult males and older female being more found with fatty change in liver. These findings are expected to have implications in the clinical and lifestyle management of NAFLD and future research.

## Data Availability

All data produced in the present study are available upon reasonable request to the corresponding author

## REFERENCES

1. European Association for the Study of the Liver (EASL); European Association for the Study of Diabetes (EASD); European Association for the Study of Obesity (EASO). EASL-EASD-EASO Clinical Practice Guidelines for the management of non-alcoholic fatty liver disease. J Hepatol. 2016; 64:1388–402. doi: 10.1016/j.jhep.2015.11.004

2. Saadeh S, Younossi ZM, Remer EM, et al. The utility of radiological imaging in nonalcoholic fatty liver disease. Gastroenterology. 2002; 123:745–50. doi: 10.1053/gast.2002.35354

3. Alam S, Fahim SM, Chowdhury MAB, et al. Prevalence and risk factors of non-alcoholic fatty liver disease in Bangladesh. JGH Open. 2018; 2:39–46. doi: 10.1002/jgh3.12044.

4. Younossi ZM, Koenig AB, Abdelatif D, et al. Global epidemiology of nonalcoholic fatty liver disease-Meta-analytic assessment of prevalence, incidence, and outcomes. Hepatology. 2016; 64:73–84. doi: 10.1002/hep.28431.

5. Dorostghol M, Gharibvand MM, Hanafi MG, et al. Comparison of size of the liver between patients with non-alcoholic fatty liver disease and healthy controls. J Family Med Prim Care. 2024; 13:425–30. doi: 10.4103/jfmpc.jfmpc_94_23

6. Joish UK, Abhishek S, Gund S. “The Plane of Y0”: Evaluating reliability of a novel method of measuring the liver with sonography using internal references. J Diag Med Sonogr 2022; 38:540–7. doi: 10.1177/87564793221092788

7. Riestra-Candelaria BL, Rodriguez-Mojica W, Jorge JC. Anatomical criteria to measure the adult right liver lobe by ultrasound. Sonography. 2018; 5:181–6. doi: 10.1002/sono.12162

8. Gan L, Chitturi S, Farrell GC. Mechanisms and implications of age-related changes in the liver: nonalcoholic Fatty liver disease in the elderly. Curr Gerontol Geriatr Res. 2011; 2011:831536. doi: 10.1155/2011/831536

